# From Community Acquired Pneumonia to COVID-19: A Deep Learning Based Method for Quantitative Analysis of COVID-19 on thick-section CT Scans

**DOI:** 10.1101/2020.04.17.20070219

**Authors:** Zhang Li, Zheng Zhong, Yang Li, Tianyu Zhang, Liangxin Gao, Dakai Jin, Yue Sun, Xianghua Ye, Li Yu, Zheyu Hu, Jing Xiao, Lingyun Huang, Yuling Tang

## Abstract

**Background:** Thick-section CT scanners are more affordable for the developing countries. Considering the widely spread COVID-19, it is of great benefit to develop an automated and accurate system for quantification of COVID-19 associated lung abnormalities using thick-section chest CT images.

**Purpose:** To develop a fully automated AI system to quantitatively assess the disease severity and disease progression using thick-section chest CT images.

**Materials and Methods:** In this retrospective study, a deep learning based system was developed to automatically segment and quantify the COVID-19 infected lung regions on thick-section chest CT images. 531 thick-section CT scans from 204 patients diagnosed with COVID-19 were collected from one appointed COVID-19 hospital from 23 January 2020 to 12 February 2020. The lung abnormalities were first segmented by a deep learning model. To assess the disease severity (non-severe or severe) and the progression, two imaging bio-markers were automatically computed, i.e., the portion of infection (POI) and the average infection HU (iHU). The performance of lung abnormality segmentation was examined using Dice coefficient, while the assessment of disease severity and the disease progression were evaluated using the area under the receiver operating characteristic curve (AUC) and the Cohen’s kappa statistic, respectively.

**Results:** Dice coefficient between the segmentation of the AI system and the manual delineations of two experienced radiologists for the COVID-19 infected lung abnormalities were 0.74±0.28 and 0.76±0.29, respectively, which were close to the inter-observer agreement, i.e., 0.79±0.25. The computed two imaging bio-markers can distinguish between the severe and non-severe stages with an AUC of 0.9680 (p-value< 0.001). *Very good* agreement (*κ* = 0.8220) between the AI system and the radiologists were achieved on evaluating the changes of infection volumes.

**Conclusions:** A deep learning based AI system built on the thick-section CT imaging can accurately quantify the COVID-19 associated lung abnormalities, assess the disease severity and its progressions.

**Key Results:** A deep learning based AI system was able to accurately segment the infected lung regions by COVID-19 using the thick-section CT scans (Dice coefficient ≥ 0.74).

The computed imaging bio-markers were able to distinguish between the non-severe and severe COVID-19 stages (area under the receiver operating characteristic curve 0.968).

The infection volume changes computed by the AI system was able to assess the COVID-19 progression (Cohen’s kappa 0.8220).

**Summary Statement:** A deep learning based AI system built on the thick-section CT imaging can accurately quantify the COVID-19 infected lung regions, assess patients disease severity and their disease progressions.

## 1 Introduction

Coronavirus Disease 2019 (COVID-19) has rapidly spread all over the world since the end of 2019, and 1, 436, 198 cases have been confirmed as COVID-19 to date (9 April 2020) [1]. Reverse-transcription polymerase chain reaction (RT-PCR) is used as the standard diagnostic method. However, it suffers from low sensitivities as report in [2, 3]. Computed tomography (CT) imaging is often adopted to confirm the COVID-19 in China and some European countries, e.g., Netherlands. CT plays a key role in the diagnosis and treatment assessment of COVID-19 due to its high sensitivity [2, 4].

The explosive growing number of COVID-19 patients requires the automated AI-based computer aided diagnosis (CAD) systems that can accurately and objectively detect the disease infected lung regions, assess the severity and the progressions. Recently, several deep learning based AI systems were developed to differentiate the COVID-19 and community acquired pneumonia (CAP) [5] or other viral pneumonia [6, 7], and to quantify the infection regions[8, 9, 10, 11]. However, all these previous AI systems built upon the high resolution thin-section CT images, which have high radiation doses and require higher costs. In contrast, the thick-section CT images from affordable CT scanners has relatively low radiation doses and are popularly used in hospitals worldwide, especially in primary care. Hence, it is worthwhile to develop an AI-based CAD system using the thick-section CT images.

In this study, we developed a fully automated AI system to quantify COVID-19 associated lung abnormalities, assess the disease severity and the disease progressions using thick-section chest CT images. Specifically, the lung and infection regions were first segmented by a deep learning based model, where the labels came from another multi-center annotated CAP CT dataset knowing that COVID-19 shares similar abnormal lung patterns with other pneumonia such as ground glass opacity (GGO), consolidation, bilateral infiltration, etc. Using the lung and infection segmentation masks, we computed the portion of infection (POI) and the average infection HU (iHU) as two imaging bio-markers, which were applied to distinguish the COVID-19 severity. Moreover, the changes of POI and iHU in patient’s longitudinal CT scans were calculated to evaluate the COVID-19 progression. For evaluation, the AI based lung abnormalities segmentation was compared to two experienced radiologists manually delineations, while the AI based assessment of disease severity and progression was compared to patients diagnosis status extracted from clinical and radiology reports. To the best of our knowledge, this is the first AI-based study to quantitatively assess the COVID-19 severity and disease progression using the thick-section CT images.

## 2 Materials and Methods

### 2.1 Patients

This study was approved by the Ethics of Committees of the First Hospital of Changsha, Hunan, China. Informed consent for this retrospective study was waived. 548 CT scans from 204 patients diagnosed with COVID-19 (RT-PCR test positive) were retrospectively reviewed for the period from 23 January 2020 to 12 February 2020 in the First Hospital of Changsha, which is the only appointed hospital healing COVID-19 patients in Changsha city, Hunan province, China. Eight patients under 18 years old were excluded for this study. The characteristics of the rest 196 adult patients were summarized in Table 1. According to the guideline of 2019-nCoV (trial version 7) issued by the China National Health Commission[12], the severity of COVID-19 includes mild, common, severe and critical types. Since there were few mild and critical cases, we categorized all the CT scans into severe group (including severe and critical) and non-severe group (mild and common). In total, we had 79 severe CT scans from 32 patients, and 452 general CT scans from 164 patients. It should be noticed that some patients were in non-severe phase when they entered the hospital, but may develop into severe phase during treatment. All the COVID-19 patients were used to test the AI system performance.

**Table 1.** Characteristics of COVID-2019 patients in this study

|  | All patients | Severe patients | Non-severe patients | p value |
| --- | --- | --- | --- | --- |
| **No.** | 196 | 32 | 164 | - |
| **Age** | 47±15 | 56±14 | 45±14 | <0.001 |
| **Male** | 96(49%) | 14(44%) | 82(50%) | 0.52 |
| **Exams** | 531 | 79 | 452 | - |
| **Patients with multiple exams** | 162(83%) | 31(97%) | 131(80%) | - |

Note: Values in parentheses are the percentage. Ages are reported as mean ± standard deviation. COVID-2019 = coronavirus disease 2019.

To train the lung abnormalities segmentation deep learning model, another multi-center pnumonia dataset was collected consisting of 558 CT scans with manual annotations. The informed consent waiver of the training data were approved by the Ethics of Committees of multiple institutes.

### 2.2 CT Protocol

All COVID-19 patients underwent the CT scanning using the GE Brivo CT325 scanner (General Electric, Illinois, the United States). The scanning protocol was as follows: 120 kV; adaptive tube current (30mAs-70mAs); pitch= 0.99-1.22 mm; slice thickness= 10 mm; field of view: 350 mm^2^; matrix, 512×512; and breath hold at full inspiration. CT images were reconstructed with 5mm slice thickness and the soft reconstruction kernel. Note that the radiation dose (3.43mGy) from the thick-section CT imaging are reasonably lower than the conventional high resolution chest CT imaging (6.03mGy, Siemens SOMATOM go.Top).

For the multi-center pneumonia dataset, the 558 CT scans were from Siemens, Hitachi, GE, Philips and United Imaging scanners. Slice thickness ranged from 1.0 - 5.0 mm. Details of the CT imaging protocols for this multi-center pneumonia dataset is presented in the Table of the supplemental material.

### 2.3 Deep Learning Model for Lung Abnormality Segmentation

We developed a 2.5D based deep learning model to segment the pneumonia infection regions using the UNet [13] structure equipped with the Resnet 34 backbone [14]. It is able to integrate the high resolution information in the axial view with the coarse continuity information along the vertical view. We also trained a standard 2D UNet to segment the lung fields in thick-section CT scans. Details of deep learning model learning is presented in the supplemental material.

### 2.4 Imaging Bio-markers Computation

Based on the lung field and infection region segmentation masks, we computed the quantitative imaging bio-markers for COVID-19, i.e., the portion of infection (POI) and the average infection HU (iHU). Specifically, we computed the POI as the infection volume divided by the total lung volume in physical unit, and the iHU as the average HU values in the infection regions.

The computed POI and iHU are consistent with latest version (the seventh) of COVID-19 diagnostic guideline released by the National Health Commission of China [12]. The guideline states that the POI is one of the principles to differentiate the severe and non-severe patients. It also reports that lung findings in chest CT may start from small subpleural GGO to crazy paving pattern and consolidation when patients conditions getting worse, which correspond to the increase in iHU changes.

### 2.5 Statistical Analysis and Evaluation Metrics

Statistical analysis was performed by SAS (version 9.4) and Matlab (version 2018b). Sensitivity and specificity were calculated using specific cutoffs by using the Youden index generated from the receiver operating characteristic curve (ROC). Cohen’s kappa statistic was used to measure of agreement between the disease progress assessment from AI and radiologists. *χ*^2^ test was used to compare differences among different groups. A two-sided p value less than 0.05 was considered to be statistically significant. The Dice coefficient was computed to evaluate agreement between the automatic infection region segmentation and the manual infection delineations by radiologists.

## 3 Results

### 3.1 Segmentation of Lung Infection Region

Examples of the infection region segmentation for severe and non-severe patients in CT were shown in Figure 1 and Figure2. To quantitatively evaluate the accuracy of segmentation, two radiologists with 20 and 15 years experiences (Z.Z and X.Y), who were blind to each other, manually delineated the infection regions of interests (ROIs) to serve as the reference standard. We randomly selected 30 CT scans of 30 patients (3 severe and 27 non-severe) and quantitatively evaluated the accuracy of the infection region segmentation on this subset. The average Dice coefficient between our method and two radiologists were 0.74±0.28 (median=0.79) and 0.76±0.29 (median=0.84), respectively. The inter-observer variability between the two radiologists was also assessed using Dice coefficient, which is 0.79±0.25(media=0.85).

**Figure 1.**
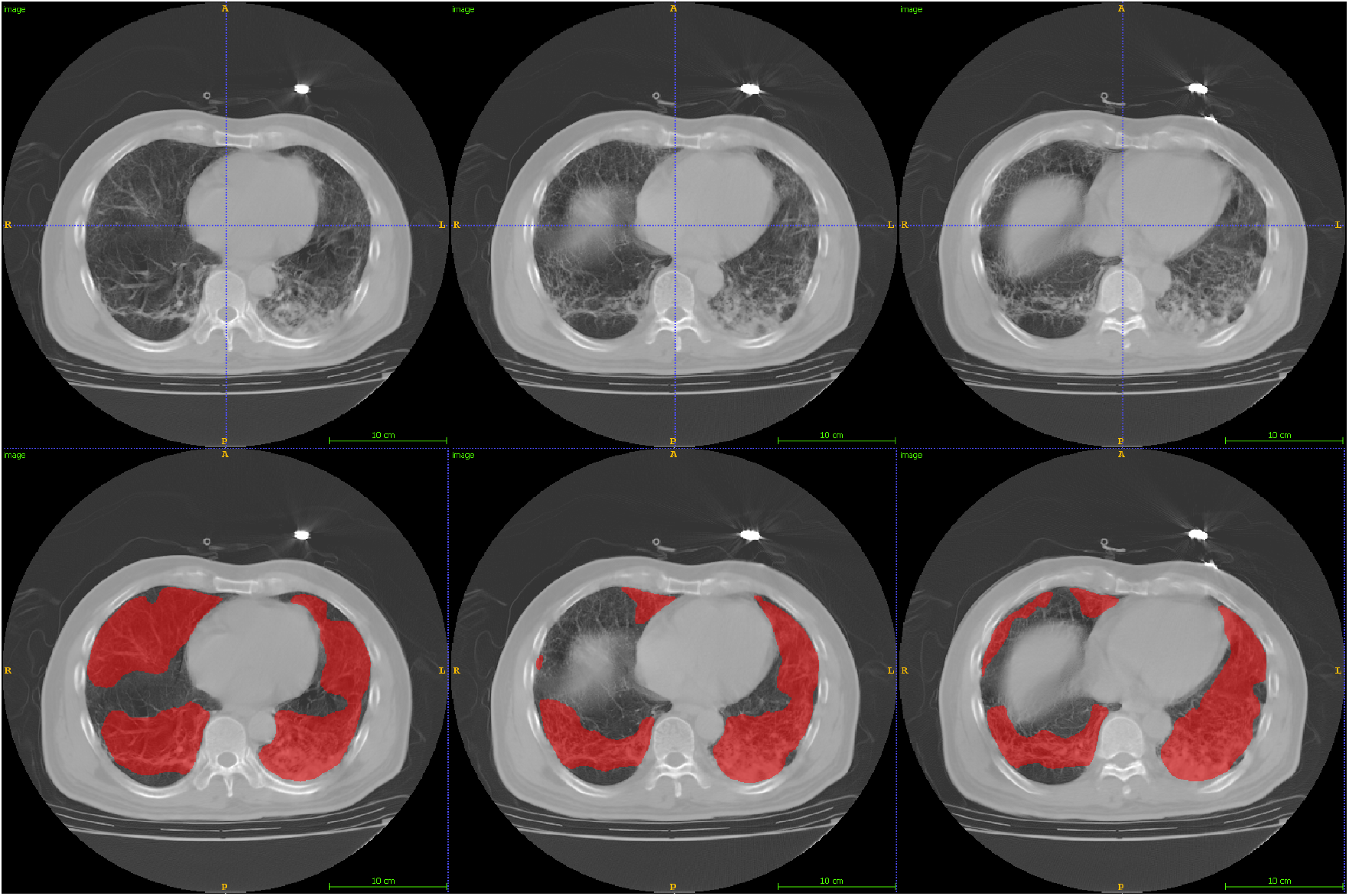
Lesion segmentation for three consecutive axial CTs from a severe patient.First row: original image; Second row: lesion segmentation image.

**Figure 2.**
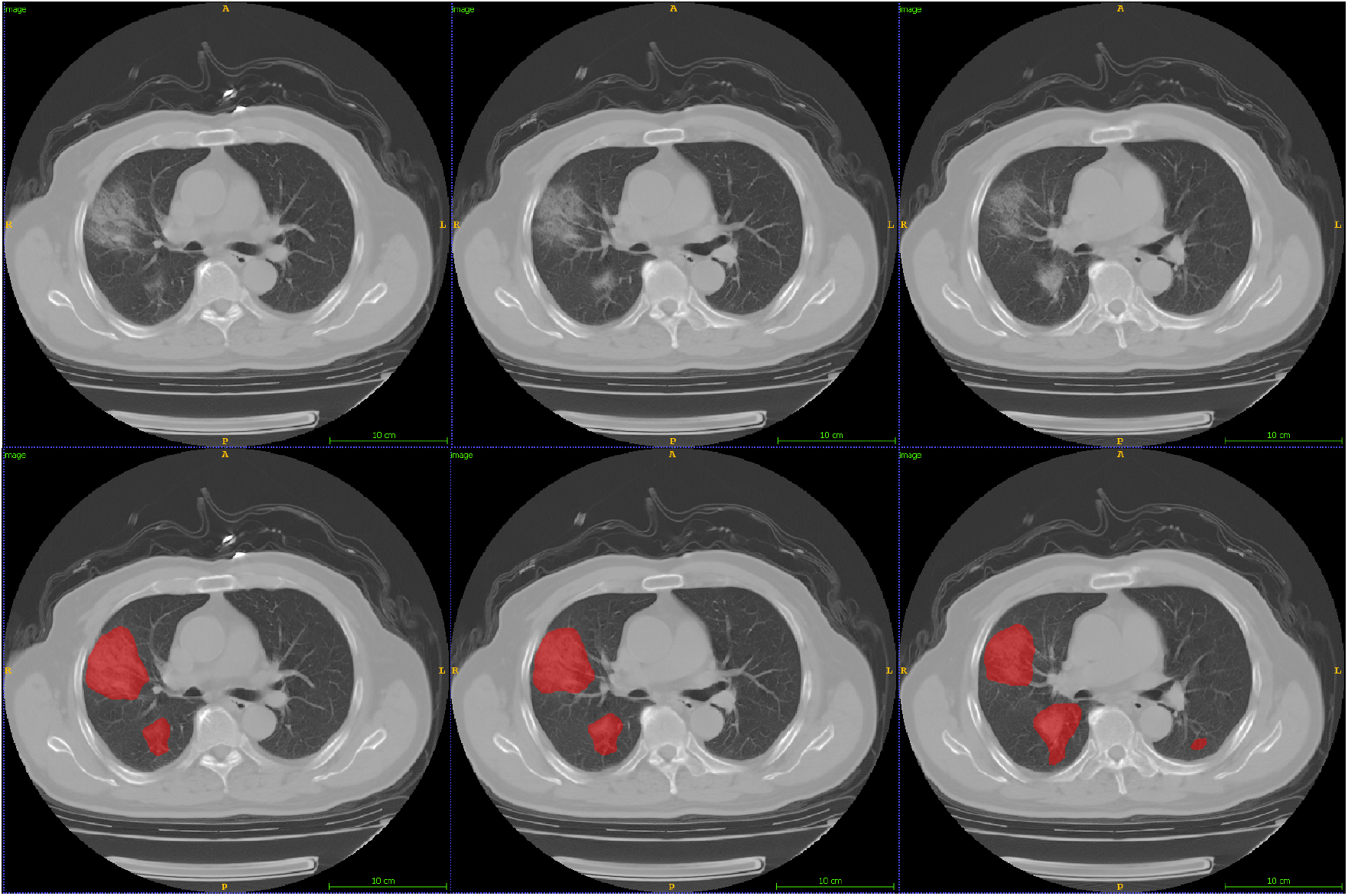
Lesion segmentation for three consecutive axial CTs from a non-severe patient.First row: original image; Second row: lesion segmentation image.

### 3.2 Assessment of Severe and Non-severe COVID-19

Based on the clinical diagnosis reports, 79 CT scans had been identified to belong to the severe group, while 452 scans were in the non-severe group. Figure 3 shows the box-plot of the computed POI and iHU for severe and non-severe groups. Note that both the POI and iHU show significant difference between severe and non-severe groups with p-value < 0.001.

**Figure 3.**
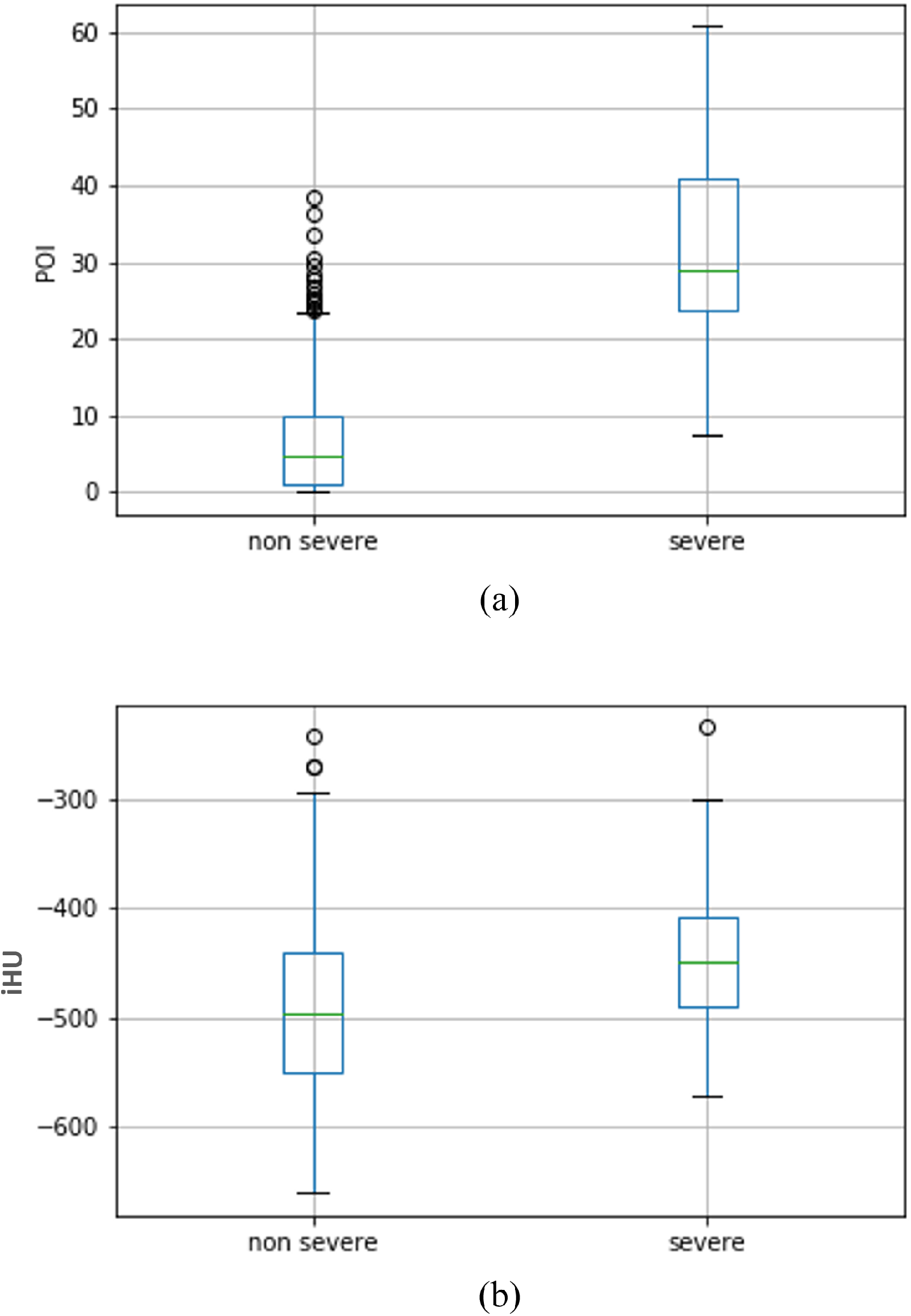
Box-plot of (a) POI and (b) iHU for the severe and non-severe patients. POI = portion of infection, iHU = infection HU.

Predictive probabilities were generated using the logistic regression model. Comparisons of different imaging bio-markers for assessment of severe and non-severe exams are shown in Table 2. Using the POI as input, the sensitivity and specificity for identifying the severe group are 92.4% 90.5%, respectively. Using the iHU as input, the sensitivity and specificity for identifying the severe group are 91.1% and 41.6%, respectively. When combining the POI with iHU, the sensitivity and specificity for identifying the severe group are 93.7% and 88.1%, respectively. The ROC curves are shown in Figure 4. The corresponding AUC values for using the iHU, POI and POI+iHU are 0.687, 0.968 and 0.968, respectively. The odds of severity at 1sd increase of POI was 18.762 [95% CI, 10.056, 35.000] (p<0.001) times higher than the baseline POI; the odds of severity at 1sd increase of iHU was 1.824 [95% CI, 1.430, 2.326] (p<0.001) times higher than the odds of severity at baseline iHU. The Akaike information criterion (AIC) for POI, iHU and POI+iHU are 174.877, 426.160 and 173.767, respectively.

**Table 2.**
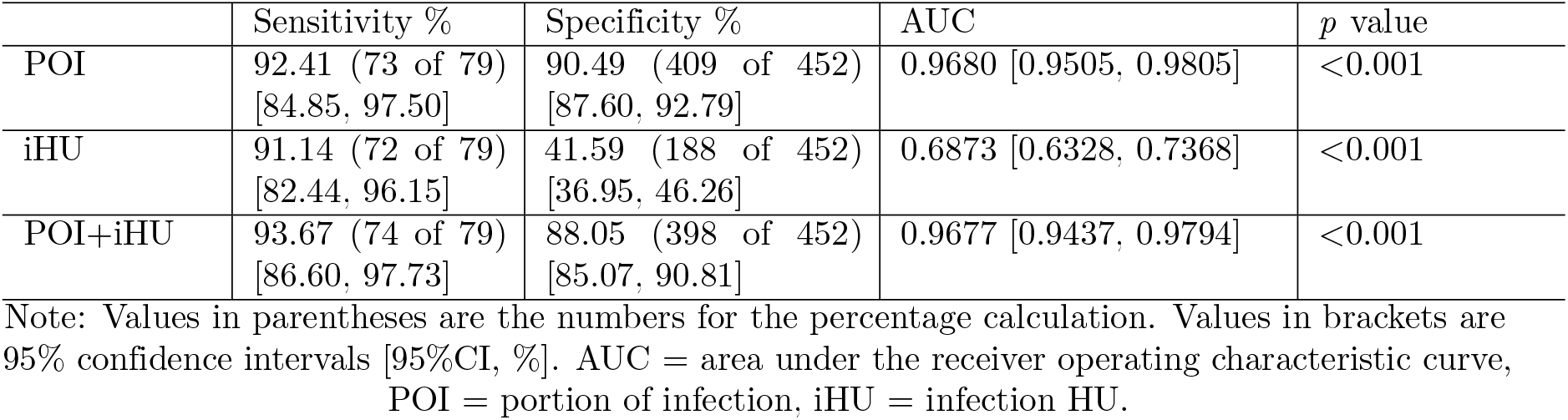
Comparisons of different imaging bio-markers for assessment of severe and non-severe exams

|  | Sensitivity % | Specificity % | AUC | p value |
| --- | --- | --- | --- | --- |
| POI | 92.41 (73 of 79)<br>[84.85, 97.50] | 90.49 (409 of 452)<br>[87.60, 92.79] | 0.9680 [0.9505, 0.9805] | <0.001 |
| iHU | 91.14 (72 of 79)<br>[82.44, 96.15] | 41.59 (188 of 452)<br>[36.95, 46.26] | 0.6873 [0.6328, 0.7368] | <0.001 |
| POI+iHU | 93.67 (74 of 79)<br>[86.60, 97.73] | 88.05 (398 of 452)<br>[85.07, 90.81] | 0.9677 [0.9437, 0.9794] | <0.001 |

Note: Values in parentheses are the numbers for the percentage calculation. Values in brackets are 95% confidence intervals [95%CI, %]. AUC = area under the receiver operating characteristic curve, POI = portion of infection, iHU = infection HU.

**Figure 4.**
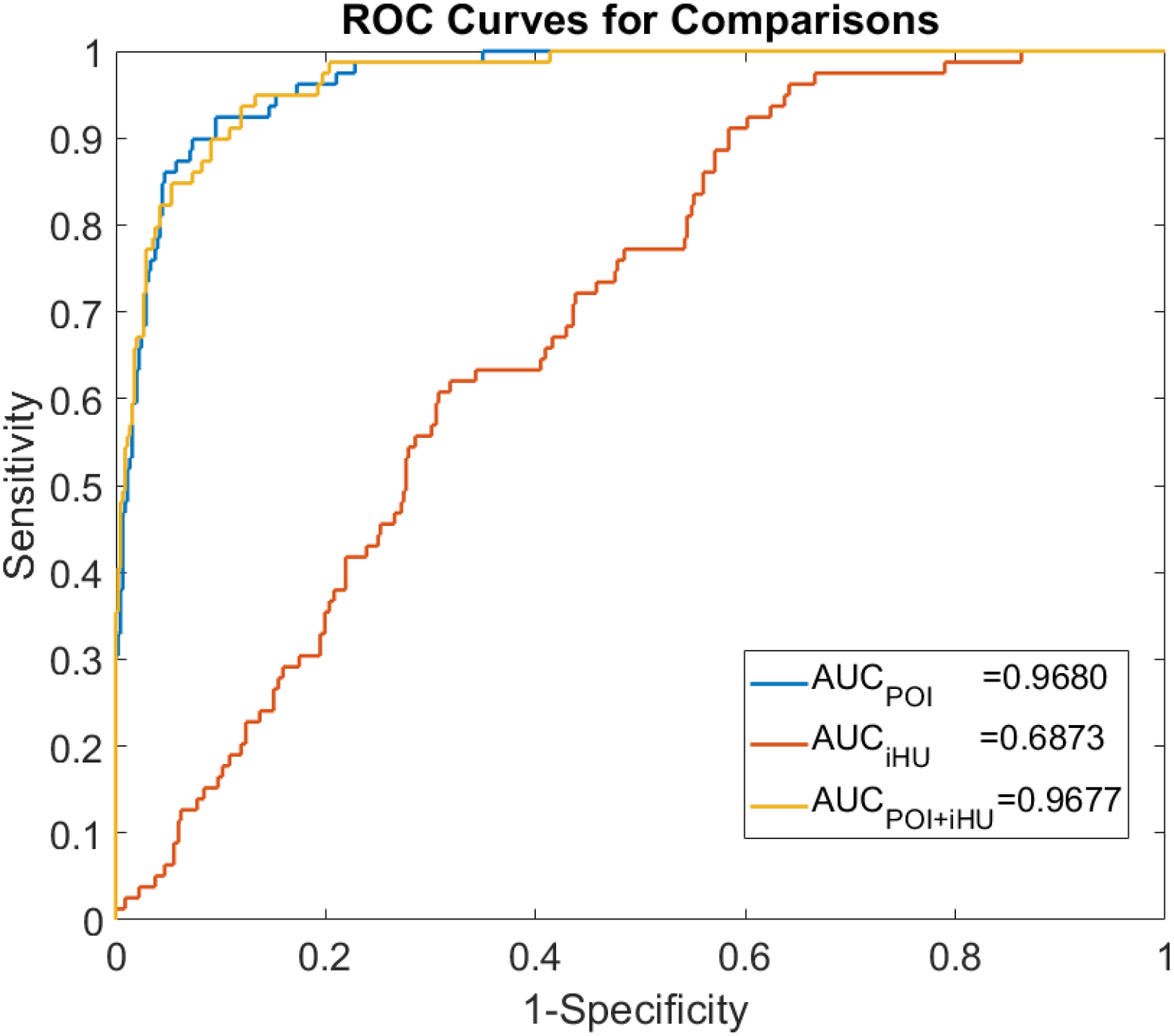
Receiver operating characteristic (ROC) curves from the logistic regression model. AUC = area under the receiver operating characteristic curve, POI = portion of infection, iHU = infection HU.

### 3.3 Assessment of Disease Progression

Figure 5 showed a qualitative example of the automatically segmented infection regions of a severe patient’s longitudinal CT scans. We calculated the changes of the POI and iHU for each consecutive CT scan pair of the patients. The key phrases extracted from patients radiology reports were used as ground-truth reference. The correspondence of the computed bio-markers changes with radiologists assessment was described in Table 3.

**Table 3.** Correspondence between the imaging bio-markers changes and radiology reports

| Phrase from radiology reports | imaging bio-markers changes | No.of phrase occurrences |
| --- | --- | --- |
| 'infection region has (partially) expanded' | increasing of POI | 86 |
| 'infection region has (partially) contracted' | decreasing of POI | 98 |
| 'Density of infection region has (partially) increased' | increasing of iHU | 43 |
| 'Density of infection region has (partially) decreased' | decreasing of iHU | 39 |

Note: Opposite phrases (partially expanded and partially contracted) that exists in six patients were excluded in this table. POI = portion of infection, iHU = infection HU.

**Figure 5.**
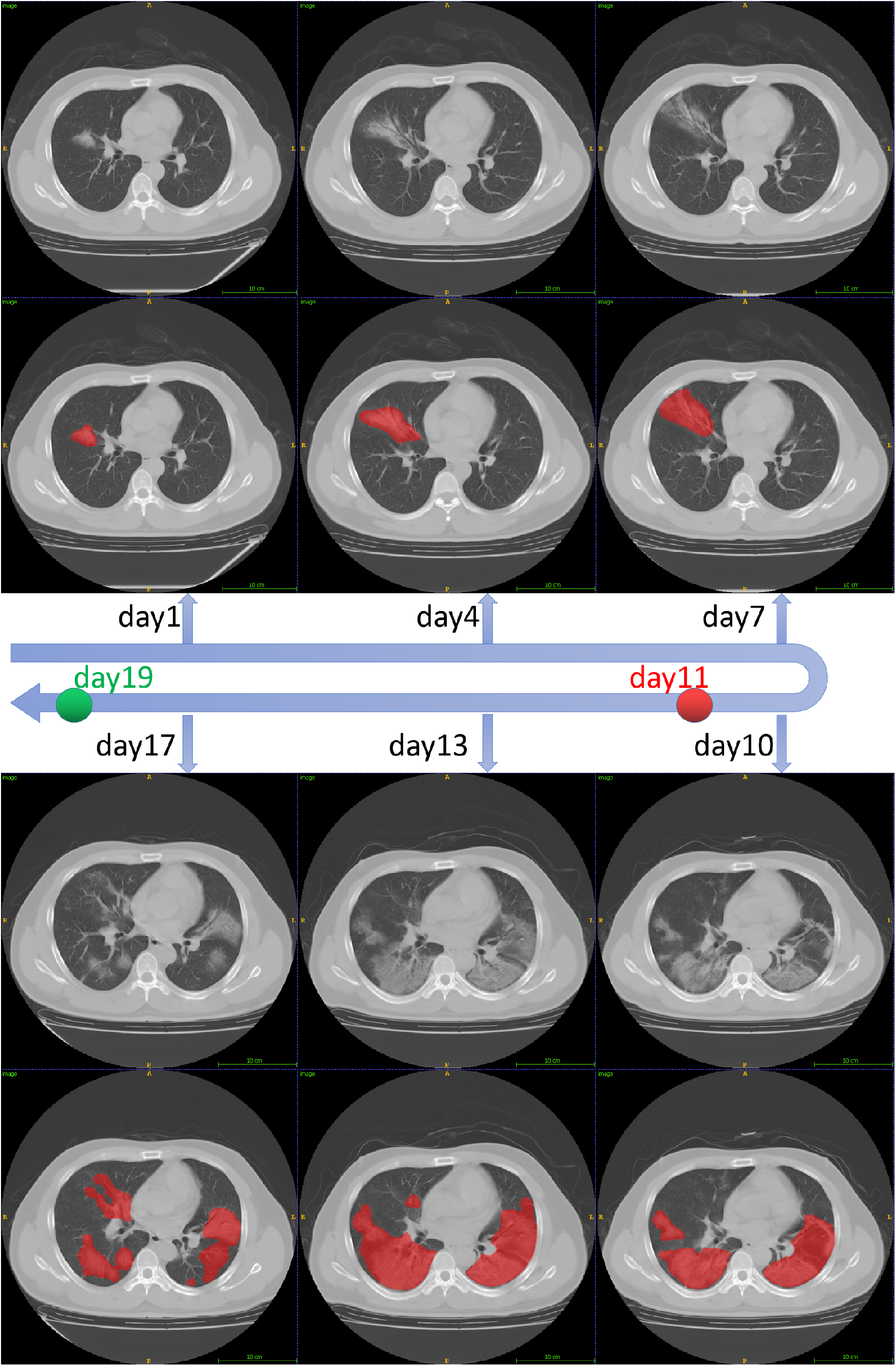
The lesion segmentation of six adjacent CT scans that taken from Jan.27 to Feb.12 for a severe patient. The red dot corresponds to the time for given the ‘severe’ diagnosis and the green point corresponds to the time for given the ‘non-severe’ diagnosis.

**Figure 6.**
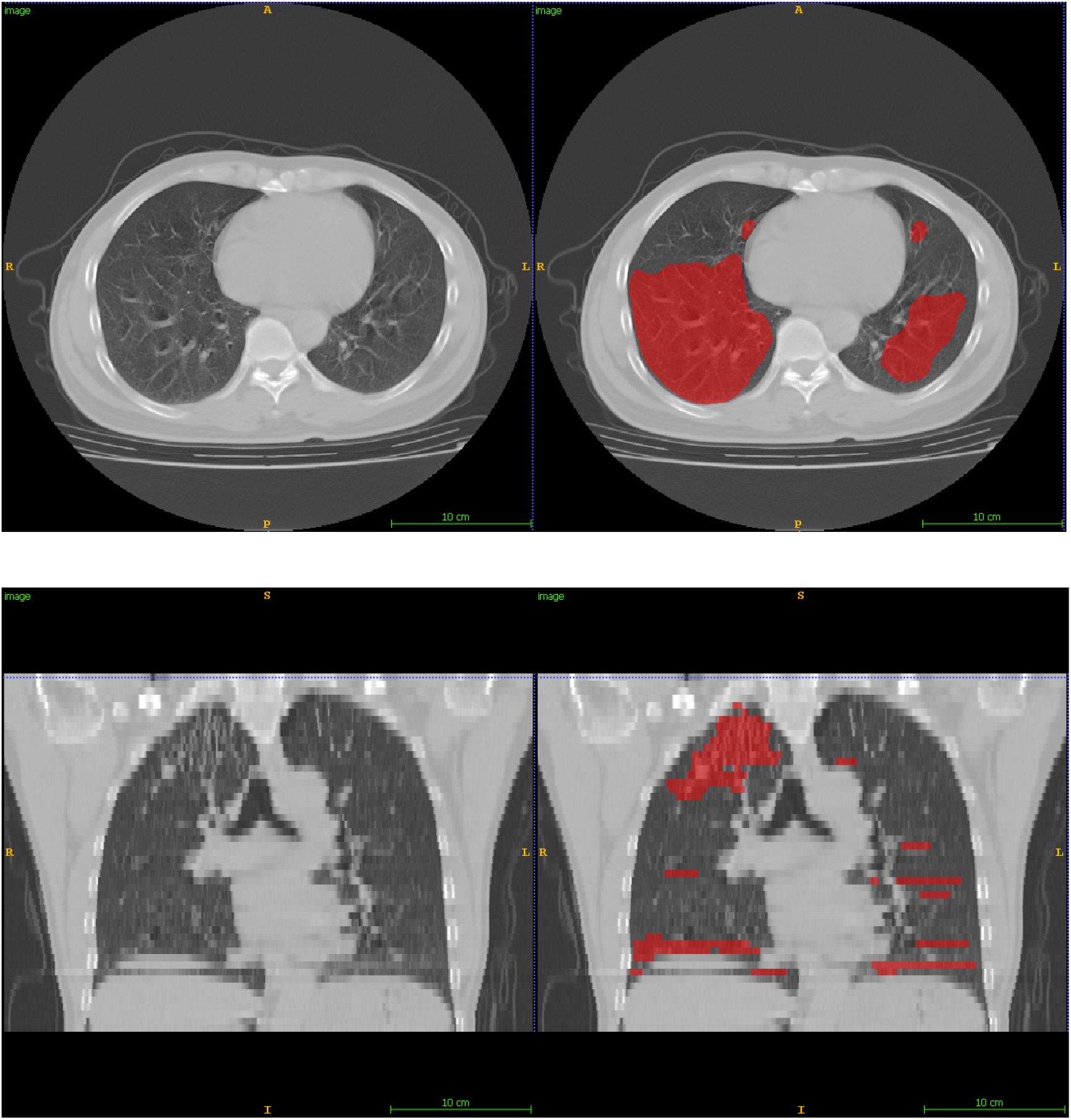
The false positive segmentation from a exam with motion artifacts.

To measure the agreement between the AI computed imaging bio-markers changes and the radiologists assessment, we first binarize the bio-markers changes. The value 1 (or 0) represented the increasing (or decreasing) of bio-markers and its corresponded phrases of radiology reports. Cohen’s Kappa was then used to measure the agreement, and the results were shown in Table 4.The *very good* and *moderate* agreement were achieved between two AI imaging bio-markers and radiologists assessment if we only consider the changes on whole lung level (ignoring the cases with phrase of ‘partially changes’). The change of POI showed overall better agreement (*very good* and *good*) with radiologists assessment than iHU (*moderate* and *fair*).

**Table 4.** Correspondence between the imaging bio-markers changes and radiology reports

|  | No.of cases | Observed agreement | Cohen's kappa | Kappa error | Strength of agreement |
| --- | --- | --- | --- | --- | --- |
| Infection region change | 142 | 0.9155 | 0.8220 | 0.0492 | very good |
| Infection region changes (including partially changes) | 184 | 0.8533 | 0.7044 | 0.0525 | good |
| Intensity changes | 54 | 0.7321 | 0.4643 | 0.1184 | moderate |
| Intensity changes (including partially changes) | 82 | 0.6829 | 0.3718 | 0.1018 | fair |

Note: Strength of agreement: 0-0.20,poor; 0.21-0.40, fair; 0.41-0.60, moderate; 0.61-0.80, good; 0.81-1.00, very good.

## 4 Discussion

In this study, we developed and evaluated an AI system for quantitative analysis of coronavirus disease 2019 (COVID-19) from thick-section chest CT scans. Our findings can be summarized as follows: 1) The deep learning model that trained on a multi-center CAP CT dataset could be directly applied for segmenting the lung abnormalities in COVID-19 patients; 2) the portion of infection (POI) and the average infection HU (iHU), with the area under the receiver operating characteristic curve (AUC) of 0.968 [95% CI: 0.951, 0.981] and 0.687 [95% CI: 0.633, 0.737], showed significant difference (p-value<0.001) in severe and non-severe COVID-19 states; 3) POI showed *Very good* agreement (*κ* = 0.8220) with the radiologist reports on evaluating the changes of infection volumes on the whole lung level.

Though high resolution CT is shown to have high sensitivity in detection of COVID-19, both cost and radiation doses are relatively high. In contrast, our study for the first time shows that an AI system can efficiently segment and quantify the COVID-19 lung infections in thick-section CT images (with relatively low radiation doses). This would benefit the developing or low-income countries, where the quantification of COVID-19 severity and the triage can be determined effectively using thick-section CT volumes of affordable CT scanners.

Our diagnosis system is an multi-stage AI system. The key step is to extract the infection regions. It is interesting that this processing modules are trained using CAP cases while the detection and segmentation accuracy is still closed to radiologist-level. Dice coefficient between the COVID-19 infected region segmentation of the AI system and two experienced radiologists were 0.74±0.28 and 0.76±0.29, respectively, which were close to the inter-observer agreement, i.e., 0.79±0.25.

Among our computed imaging bio-markers, only the POI shows high sensitivity and specificity for differentiating the severe from non-severe COVID-19 groups. This indicates that the POI is an effective imaging bio-marker to assess the severity of COVID-19 patients. Although the iHU value is also able to reflect infection progress, however, it is affected by several other disease irrelevant factors, such as the reconstruction slice thickness and the respiration status[15, 16]. For instance, consolidation on HRCT images might be displayed as GGO on thick-section CT images.

The changes of volume and density of the infection region are two key indicators that used by radiologists for COVID-19 progression assessment. However, it is time consuming (or even impractical) for radiologists to produce the quantitative measurements for this longitudinal analysis. Our AI system provides a quantitative and objective measurement, i.e., the POI, which shows strong agreement with radiologist qualitative judgements. More importantly, the AI based longitudinal disease quantification is precise, reproducible and fast, which can reduce the reading time of radiologists for COVID-19 each patient and improve the quality of the disease progression assessment[10].

This study has several limitations. Firstly, we only evaluated changes of imaging bio-markers at the whole lung level in certain phrase. Although our model can compute the bio-markers at the lobe level, the standard phrases from the radiology reports were mostly at the whole lung level. Furthermore, some phrases in the reports like ‘lesion absorption’ might respond to either infection region decreasing or HU value reduction. Thus it needs more sophisticated and precise analysis evaluating our model in the future. Secondly, motion artifacts due to respiration and heart motion may cause false positive segmentation in the AI system. We noticed that some false positive segmentation affected the longitudinal infection evaluations6. One possible solution is to identity the motion artifacts before applying the infection segmenting. Finally, our model was only tested the COVID-19 positive patients. A recent study has shown that a deep learning based AI classification model can detect the COVID-19 and distinguish it from the community acquired pneumonia and other non-pneumonic lung diseases using thin-section HRCT [5]. As the next step, it would be interesting to see if our model can also differentiate the pneumonia caused by COVID-19 and other factors using the thick-section CT imaging.

In conclusion, a deep learning based AI system is developed to quantify COVID-19 abnormal lung patterns, assess the disease severity and the progression using thick-section chest CT images. The imaging bio-makers computed from the AI system could be used for reproducing several findings of infection change from the reports by radiologists. These results demonstrate that the deep learning based tool has the ability to help radiologists on diagnosing and follow-up treatment for COVID-19 patients based on CT scans.

## Data Availability

Not applicable

## Supporting Information

This work was partially supported by National Natural Science Funding of China (No.61801491), Natural Science Funding of Hunan Province (No.2019JJ50728) and The Research Program of the Hunan Health and Family Planning Commission(No.B20180393).

## Abbreviations

AUC: area under the receiver operating characteristic curve
CI: confidence interval
COVID-19: coronavirus disease 2019
POI: portion of infection
iHU: average infection Hounsfield unit

